# Genetically-proxied lower PAR1 and renal phenotypes: a drug-target Mendelian Randomization study

**DOI:** 10.1101/2024.08.08.24311690

**Authors:** Haotian Tang, Tom R Gaunt, Venexia M Walker

## Abstract

In clinical trials, the anti-protease-activated receptor-1 (PAR1) therapy, vorapaxar, has been tested as a treatment for nephrotic syndrome (NS). This study aimed to investigate the causal relationship of PAR1 (encoded by the gene *F2R*) with renal phenotypes using drug-target Mendelian randomization (MR).

First, we performed colocalization analyses to confirm whether PAR1/*F2R* expression instruments were shared between plasma/blood and kidney tissue. We selected PAR1/*F2R* expression instruments from the UK Biobank-Pharma Proteomics Project (UKB-PPP), eQTLGen, and Genotype-Tissue Expression (GTEx). Kidney *F2R* expression data were obtained from NephQTL2. Second, given the known experimental evidence of the vorapaxar effect on PAR1 inhibition, we conducted drug-target MR to investigate the causal effects of genetically lower PAR1 protein levels or *F2R* expression levels on thrombotic diseases as positive controls from FinnGen, including venous thromboembolism (VTE), deep venous thrombosis (DVT), and arterial thromboembolism (AET). Finally, we performed drug-target MR to investigate the causal relationship of genetically lower PAR1 protein or *F2R* expression levels from plasma/blood and kidney tissues on renal diseases and function. We included five renal phenotypes as our main outcomes: CKD, microalbuminuria (MA), NS, estimated GFR (eGFR), and urinary albumin-to-creatinine ratio (uACR). Renal genome-wide association studies (GWAS) data were obtained from the CKDGen consortium, except for NS, which was obtained by meta-analysing FinnGen and a novel GWAS in the UKB. In addition, we also used serum albumin from UKB as a further outcome in a sensitivity analysis.

Colocalization analyses identified shared genetic variants between blood and tubulointerstitial *F2R* expression (posterior probabilities are 89.1% for eQTLGen and 95.4% for GTEx) but not between plasma PAR1 and tubulointerstitial *F2R* expression. We present MR findings as *β* or odds ratios (ORs) with 95% confidence intervals (CIs) per unit decrease in plasma protein or gene expression, or per effect allele, consistent with the direction of effect when taking vorapaxar. Genetically-proxied lower PAR1 and *F2R* expression reduced risk of VTE with ORs of 0.84 [95% CI 0.71 to 1.00] (UKB-PPP), 0.94 [0.88 to 1.01] (eQTLGen) and 0.89 [0.75 to 1.06] (GTEx), indicating that genetically-instrumented lower PAR1 is directionally consistent with the effect of vorapaxar. Genetically-proxied lower PAR1 and *F2R* expression increased the risk of developing CKD (UKB-PPP: OR=1.17 [0.99 to 1.38]; eQTLGen: OR=1.12 [1.02 to 1.23]; GTEx: OR=1.24 [1.05 to 1.46]), but may decrease eGFR (eQTLGen: *β* =-0.02 [-0.06 to 0.01], GTEx: *β* =-0.04 [-0.08 to 0.01], UKB-PPP: *β* =-0.01, [-0.05, 0.04]). The effect of genetically-proxied PAR1 on NS was inconclusive due to lack of power.

Our study suggests that genetically-proxied lower PAR1 and *F2R* expression reduces VTE risk, and increases CKD risk, but provides less evidence on reducing eGFR or increasing uACR. Evidence regarding NS was inconclusive. For NS patients, the decision to use anti-PAR1 treatment should be made with caution, given the potential renal implications and the current lack of conclusive evidence of its impact on NS.

## Introduction

Vorapaxar has been used to prevent atherothrombotic events in patients with prior myocardial infarction, ischemic stroke, or peripheral arterial disease. Vorapaxar blocks protease-activated receptor 1 (PAR1) (encoded by *F2R* gene) activation on platelet membrane through reversible antagonism and inhibits thrombin-induced platelet aggregation and thrombin receptor agonist peptide (TRAP)-induced platelet aggregation.^1^ May et al. (2023) showed that podocyte-specific activation of PAR1 leads to early severe nephrotic syndrome in mice and there might be unknown circulating factors in patients with NS that cause relapsing focal segmental glomerulosclerosis (FSGS) after kidney transplantation.^2^ Anti-PAR1 treatments have also been studied in clinical trials for NS. However, no genetic epidemiological study has previously investigated the causal effect of PAR1 inhibition on NS.

NS is a clinical condition characterized by significant glomerular protein leakage into the urine. It is diagnosed by the excretion of more than 3.5 *g* of protein per 1.73 *m*^2^ of body surface area in 24 hours or a urinary protein-to-creatinine ratio (uPCR) exceeding 3.5 *mg/mg*.^3, 4^ The common etiologies of NS include diabetic nephropathy, minimal change disease (MCD), focal segmental glomerulosclerosis (FSGS), and membranous nephropathy. Among these, FSGS is the predominant glomerular cause of end-stage kidney disease (ESKD) in the United States,^5^ which is the last stage of chronic kidney disease (CKD). CKD is a progressive disorder characterized by the presence of kidney damage or decreased kidney function for more than 3 months. Its stages are classified by two dimensions: glomerular filtration rate (GFR) and urinary albumin-to-creatinine ratio (uACR), which indicates the severity of albuminuria.^6–8^ While there is an ongoing debate about whether uACR should replace uPCR, both measurements are effective and increases in uACR correspond to increases in uPCR.^9^ Mechanistically, the loss of albumin signifies damage to the glomerular filtration system. In contrast, the loss of various proteins, including albumin, indicates a broader spectrum of kidney dysfunction, such as tubulointerstitial damage.^10, 11^

Traditional observational research faces challenges in drawing causal inferences due to potential biases from reverse causality and confounding.^12^ Mendelian randomization (MR) addresses these limitations by using genetic variants as instrumental variables to assess causal effects. Based on Mendel’s Laws of inheritance, offspring genotypes are less likely to be associated with population confounders, such as behavioral and social factors, and reverse causation is avoided as germline genetic variants are established at conception and precede the risk factors under investigation.^13, 14^ Drug-target MR, which uses a genetic proxy for the effect of a drug target as an exposure, has been applied for drug repurposing, adverse effect assessment, and target identification of novel drugs.^15–22^

Our study aimed to investigate the causal relationship of PAR1 (encoded by the gene *F2R*) with renal phenotypes using drug-target MR. First, we performed colocalization analyses to confirm whether PAR1 instruments were shared between plasma/blood and kidney tissue. Second, given the known experimental evidence of the vorapaxar effect on PAR1 inhibition, we conducted drug-target MR to investigate the causal effects of genetically lower PAR1 protein levels or *F2R* expression levels on thrombotic diseases as positive controls, including venous thromboembolism (VTE), deep venous thrombosis (DVT), and arterial thromboembolism (AET). Finally, we performed drug-target MR to investigate the causal relationship of genetically lower PAR1 protein or *F2R* expression levels from plasma/blood and kidney tissues on renal diseases and function. We included five renal phenotypes as our main outcomes: CKD, microalbuminuria (MA), NS, estimated GFR (eGFR), and uACR. In addition, we also used serum albumin as a further outcome in a sensitivity analysis.

## Methods and materials

### Study design

This study is reported according to the “Strengthening the reporting of observational studies in epidemiology using Mendelian Randomization” (STROBE-MR) guidelines.^23^ An overview of the study design is shown in Figure 1.

**Figure 1:**
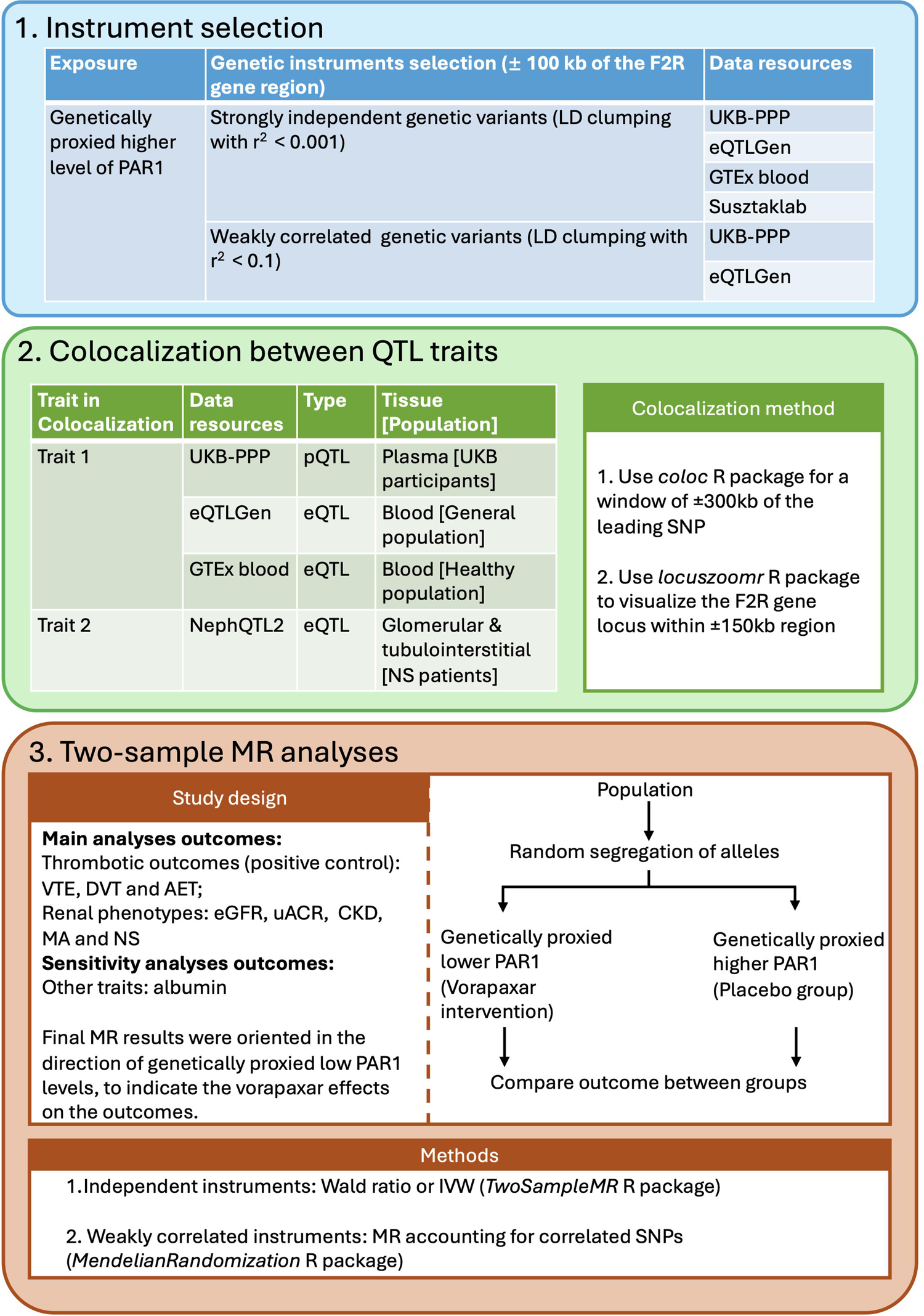
The study overview. We sequentially performed 1) LD-clumping, 2) colocalization between QTLs, and 3) drug-target MR analyses. UKB-PPP, UK Biobank Pharma Proteomics Project; eQTLGen, the eQTLGen Consortium; GTEx, The Genotype-Tissue Expression project; Susztaklab, Susztaklab Human Kidney eQTL Atlas; CKDGen, the Chronic Kidney Disease Genetics Consortium; eGFR, estimated glomerular filtration rate; NS, nephrotic syndrome; uACR, urinary albumin-creatinine ratio; MA, microalbuminuria; ATE, Arterial thromboembolism; VTE, venous thromboembolism; DVT, deep vein thrombosis; pQTL/eQTL, protein/expression quantitative trait locus; LD, linkage disequilibrium.

### Data sources

#### Exposures

We extracted genetic variants to use as instruments for the plasma PAR1 protein level from the UK Biobank Pharma Proteomics Project (UKB-PPP) (N=54,219),^24^ as well as the whole-blood *F2R* expression level from eQTLGen (N=31,684)^25^ and the Genotype-Tissue Expression (GTEx) project (v8) (N=670).^26^ We also extracted genetic variants to use as instruments from the Susztaklab for the *F2R* expression levels in a meta-analysis of kidney tissues (N=686),^27^ in glomeruli tissue (N=303) and Tubule tissue (N=356).^28^ In addition, for the meta-analyzed eQTLs from kidney tissue, Liu et al. (2022) conducted a meta-analysis of GWAS data from four non-overlapping studies, including Sheng et al., GTEx kidney tissue and NephQTL2.^29^ To simplify the description of our methods and results, we refer to plasma PAR1 levels, whole blood *F2R* expression, and kidney *F2R* expression as PAR1, blood *F2R*, and kidney *F2R* respectively. We refer to Susztaklab Kidney and Susztaklab Tubule tissue as Kidney and Tubule tissue respectively.

#### Outcomes

We obtained summary statistics from the GWAS of VTE (21,020 cases and 408,189 controls), DVT (6,501 cases and 422,708 controls), and AET (1,883 cases and 427,326 controls) from FinnGen R10^30^ to use as positive control outcomes.

For the renal phenotypes in our main analyses, we obtained summary statistics from the CKD Genetics (CKDGen) Consortium. These included CKD (41,395 cases and 439,303 controls), creatinine-based eGFR (N=567,460),^31^ uACR (N=547,361) and MA (51,861 cases and 297,093 controls).^32^ All GWAS were performed in individuals of European ancestry, except the GWAS for MA which included trans-ethnic participants. For the NS outcome, we used METAL to meta-analyze the FinnGen NS GWAS^33^ with a novel GWAS we conducted in UKB (see Supplementary Methods) (meta-analyzed NS: 1,266 cases and 887,979 controls). For our sensitivity analyses, we obtained summary statistics for serum albumin level (N=315,268) from UKB via the IEU OpenGWAS platform^34^ (ID: ukb-d-30600_irnt).

#### Data specific for colocalization

To confirm that the genetic variants of plasma protein and whole-blood *F2R* expression levels are shared with the *F2R* expression in the kidney, we obtained glomerular (N=240) and tubulointerstitial (N=311) *F2R* eQTLs from NephQTL2,^29^ where eQTL analysis used RNA samples from microdissected biopsies from the the Nephrotic Syndrome Study Network (NEPTUNE) study, an NS-disease-specific cohort.^35^ These data were only used for colocalization analyses.

#### Instrument selection

We selected instruments that were robustly associated with *F2R* expression or PAR1 protein levels at genome-wide significance (*p <* 5 *×* 10*^−^*^8^) and located within 100kb upstream and downstream of the *F2R* gene [Ensembl: ENSG00000181104; gene location: chr5:76,716,126-76,735,770 (GRCh38/hg38)]. To identify independent genetic variants, we performed linkage disequilibrium (LD) clumping with a window of 10,000kb and an LD threshold (*r*^2^) of less than 0.001.^36^ We calculated F-statistics to evaluate the strength of the genetic instruments for each MR analysis.^37–39^ F-statistics are typically interpreted using an arbitrary threshold of 10 to indicate a strong instrument.

### Main analyses

#### Colocalization between PAR1 cis-QTL and disease-specific *F2R* eQTL

To quantify the probability of a shared genetic variant between PAR1/blood *F2R* and kidney *F2R* from the NS-specific cohort and indicate if the selected instruments are valid, we conducted genetic colocalization. As the data from the Susztaklab only included a limited number of significant genetic variants, we could not conduct colocalization between Susztaklab and NEPTUNE. For each colocalization analysis, we applied the default prior probabilities for a single SNP being associated with each phenotype individually (*p*_1_ = 1 *×* 10*^−^*^4^; *p*_2_ = 1 *×* 10*^−^*^4^) and with both phenotypes simultaneously (*p*_12_ = 1 *×* 10*^−^*^5^). Colocalization considers five potential hypotheses. We were primarily interested in hypothesis 4 (H4), which assumes that there is a shared causal variant that is associated with both traits.^40^ We used the posterior probability of H4 (PP4) *>* 80% to suggest that there was a shared variant associated with both traits in the region. We used the cis-QTL summary level data from UKB-PPP, eQTLGen, and GTEx blood in turn as trait one and the eQTL summary level data from NephQTL2 as trait two.^29^

#### Drug-target MR

Given the known effect of vorapaxar on anti-platelet aggregation, we selected thrombotic diseases as positive control outcomes, including VTE, DVT, and AET. We performed drug-target MR to investigate the causal effects of genetically-proxied lower PAR1, blood *F2R*, and kidney *F2R* on thrombotic diseases to confirm whether the causal effects of genetically-proxied lower PAR1 and *F2R* are in the same direction as the vorapaxar effect.

We then conducted drug target MR analyses to explore the causal effects of genetically-proxied lower PAR1, blood *F2R*, and kidney *F2R* on renal phenotypes. To avoid bias from sample overlap in these analyses,^41^ we restricted the data included in analyses for some outcomes to remove UKB participants when using UKB-PPP data as the exposure. For example, we used summary statistics from FinnGen only for NS.

In the main drug-target MR analyses, we used the Wald ratio method^42, 43^ for single SNP instruments and the fixed-effects inverse-variance-weighted (IVW) method^44^ for multiple SNP instruments. As shown in Figure S1, MR requires three assumptions to be satisfied. They are: (IV1) The genetic instruments must be strongly associated with exposure (Relevance); (IV2) There must be no shared causes between the genetic instruments and the outcome (Independence); (IV3) The genetic instruments must be associated with the outcome via the exposure only (Exclusion restriction).^13^ The Relevance assumption is the only assumption that we can ensure is met, which we did by using instruments that are genome-wide significant for the exposure of interest. Typically alternative MR methods, such as MR Egger,^45^ should be used to examine violations of the other assumptions. However, many of our instruments contained too few SNPs for us to apply these methods.

### Sensitivity analyses

#### Genetically-proxied lower *F2R* on serum albumin

We repeated our analyses using serum albumin as an outcome as decreased serum albumin levels are associated with kidney function decline.^46^ This helped to ensure that our results were not driven by any specific renal phenotype or renal function measurement method but reflected a broader effect on renal-related outcomes.

#### MR accounting for correlated instruments

When only a few independent genetic instruments are available, MR results are sensitive to the choice of the instruments. We increased the number of genetic instruments in our MR analyses by raising the LD threshold (*r*^2^) to 0.1 for LD clumping. This resulted in weakly correlated instruments, potentially violating MR assumptions. Therefore, we performed MR analyses using IVW accounting for LD between genetic variants. These analyses used LD matrices from the same reference panel as the original LD clumping.^47^

## Results

All MR results are presented in the direction of genetically-proxied lower PAR1 to proxy the effect of vorapaxar. For continuous and binary outcomes, effect estimates are presented as the beta coefficient (*β*) or the odds ratio (OR) of the outcome per standard deviation (SD) or per effect allele decrease in exposure respectively, accompanied by a 95% confidence interval (CI).

### Instrument selection

We identified one genetic variant from UKB-PPP, three from eQTLGen, one from GTEx, one from Susztaklab Kidney, and one from Susztaklab Tubule tissue, using LD clumping with an *r*^2^ threshold of 0.001. There were no significant genetic variants from the glomeruli. The F statistics for all genetic instruments were larger than 10, indicating weak instrument bias is unlikely to have impacted our MR results. Details of each genetic instrument are provided in Supplementary Table (ST) 1.

## Main Results

### Co-localization between PAR1 cis-QTL and disease-specific *F2R* eQTL

There were no shared or distinct genetic variants from the colocalization analyses between any plasma PAR1/blood *F2R* expression and *F2R* in glomerular tissue from NEPTUNE (UKB-PPP: PP3=18.4%, PP4=5.2%; eQTLGen: PP3=15.6%, PP4=18.7%; GTEx: PP3=18.0% and PP4=9.4%).

There was no colocalization between the PAR1 pQTL from UKB-PPP and the PAR1 eQTL from tubulointerstitial tissue of NEPTUNE (PP3=42.1% and PP4=3.5%). Colocalization analyses revealed a shared genetic variant (rs1472215) between *F2R* expression in blood from eQTLGen and in tubulointerstitial tissue from NEPTUNE (PP4 = 89.1%), as well as a shared genetic variant (rs250753) between *F2R* expression in blood from GTEx and NEPTUNE (PP4 = 95.4%) (Figure 2 and ST 2).

**Figure 2:**
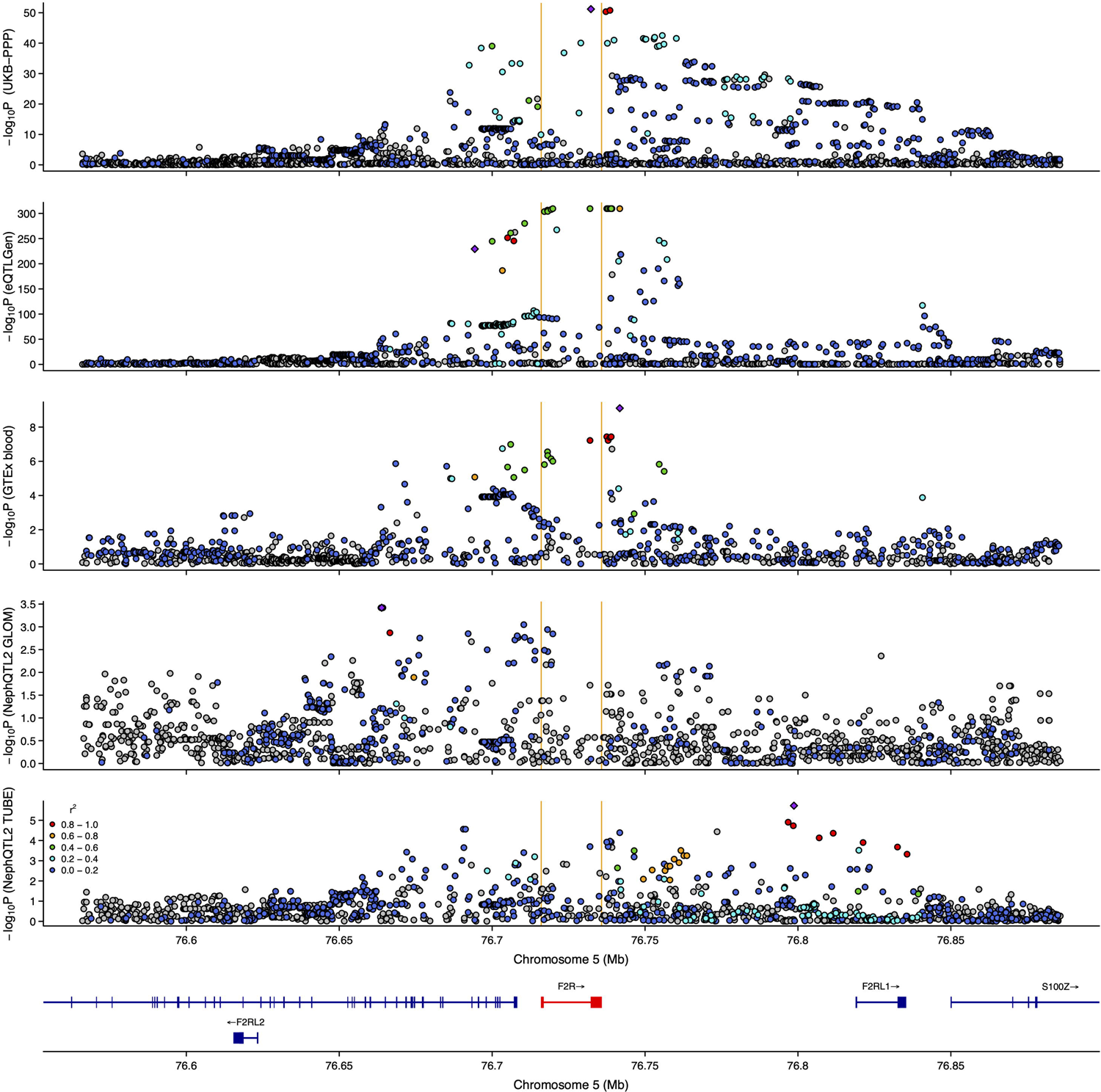
A LocusZoom plot showing genetic colocalization results between blood *F2R* gene expression, plasma PAR1 protein levels and kidney *F2R* gene expression from NephQTL2 data. Purple squared dots are the leading SNPs in each study. The Y-axis is presented as *−log*_10_*P* of SNPs with their corresponding data resources. UKB-PPP, UK Biobank Pharma Proteomics Project; eQTLGen, the eQTLGen Consortium; GTEx, The Genotype-Tissue Expression project; NephQTL2 GLOM and TUBE, glomerular and tubulointerstitial tissues from NephQTL2.

These results identified shared variants of *F2R* expression between blood from the general population and kidney tubulointerstitial tissues from the NS-specific population, indicating the instruments are likely to be valid. A lack of evidence for colocalization between blood PAR1 protein and kidney *F2R* expression may indicate the distinct mechanisms of producing protein and expressing mRNA. A lack of evidence of colocalization for glomerular tissue might reflect limited statistical power (minimum p-value=3.8 *×* 10*^−^*^4^). Consequently, we did not filter out any instruments based on the colocalization results.

### Positive control: genetically-proxied lower PAR1/*F2R* on thrombotic diseases

Genetically-proxied lower PAR1, blood, and kidney *F2R* may reduce the risk of developing VTE and DVT (Figure 3 and ST 3) (For VTE: UKB-PPP: OR=0.84, 95% CI: 0.71 to 1.00 and Tubule: OR=0.82, 95% CI: 0.71 to 0.95. For DVT: UKB-PPP: OR=0.72, 95% CI: 0.54 to 0.97). However, results for AET were inconclusive due to a lack of power (e.g. UKB-PPP: OR=1.04, 95% CI: 0.60 to 1.78).

**Figure 3:**
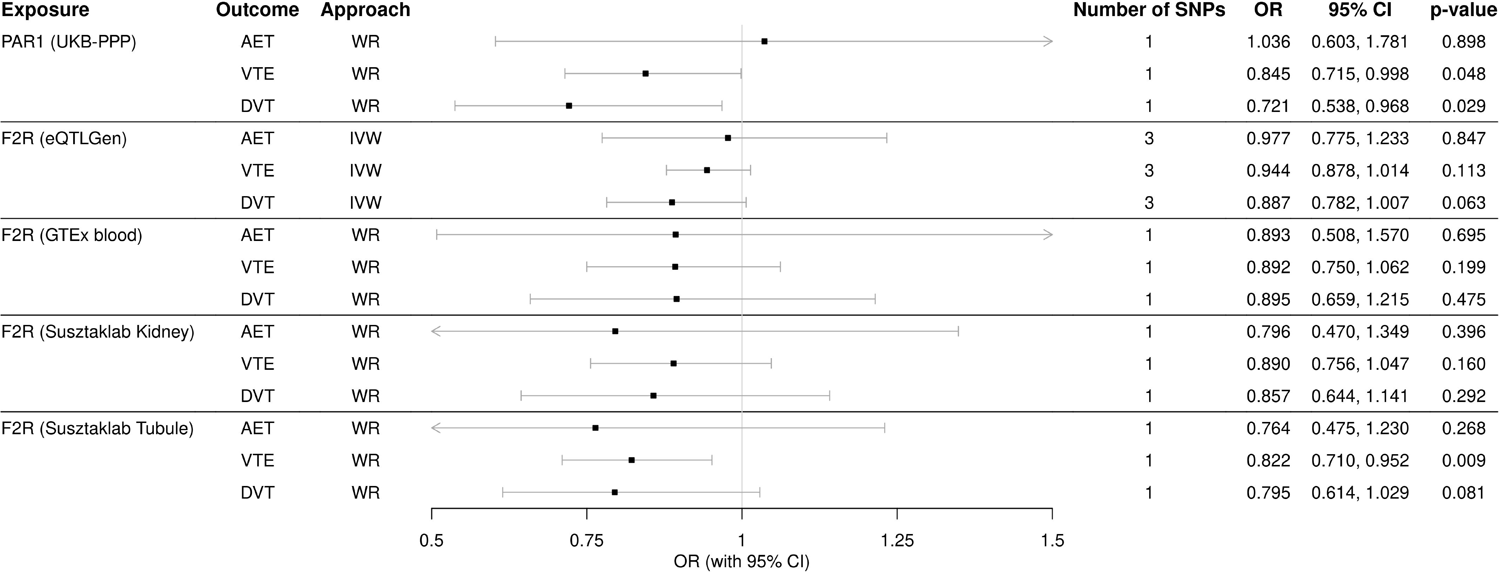
Results from MR analyses to estimate the causal effects of genetically-proxied lower PAR1 level on thrombotic diseases. UKB-PPP, UK Biobank Pharma Proteomics Project; eQTL-Gen, the eQTLGen Consortium; GTEx, The Genotype-Tissue Expression project; Susztaklab, Susztaklab Human Kidney eQTL Atlas; ATE, Arterial thromboembolism; VTE, venous throm-boembolism; DVT, deep vein thrombosis; WR, Wald ratio; IVW, inverse-variance-weighted.

### Genetically-proxied lower PAR1/*F2R* on renal phenotypes

As shown in Figure 4 and ST 4, genetically-proxied lower PAR1 and blood *F2R* might increase the risk of developing CKD (UKB-PPP: OR=1.17, 95% CI: 0.99 to 1.38; eQTLGen: OR=1.12, 95% CI: 1.02 to 1.23; GTEx: OR=1.24, 95% CI: 1.05 to 1.46). However, the causal effects of genetically-proxied lower kidney *F2R* expression were OR=0.95 (95% CI: 0.82 to 1.10) in Kidney; OR=1.07 (95% CI: 0.93 to 1.23) in Tubule. The causal effects of genetically-proxied lower blood *F2R* on MA risk were OR=0.99 (95% CI: 0.95 to 1.04) in eQTLGen; OR=1.00 (95% CI: 0.89 to 1.13) in GTEx. The causal effect of genetically-proxied lower kidney *F2R* on MA were OR=1.09 (95% CI: 0.98 to 1.22) in Kidney; OR=1.10 (95% CI: 1.00 to 1.22) in Tubule. The causal effect of genetically-proxied lower PAR1, blood, or kidney *F2R* on NS is inconclusive due to a lack of power.

**Figure 4:**
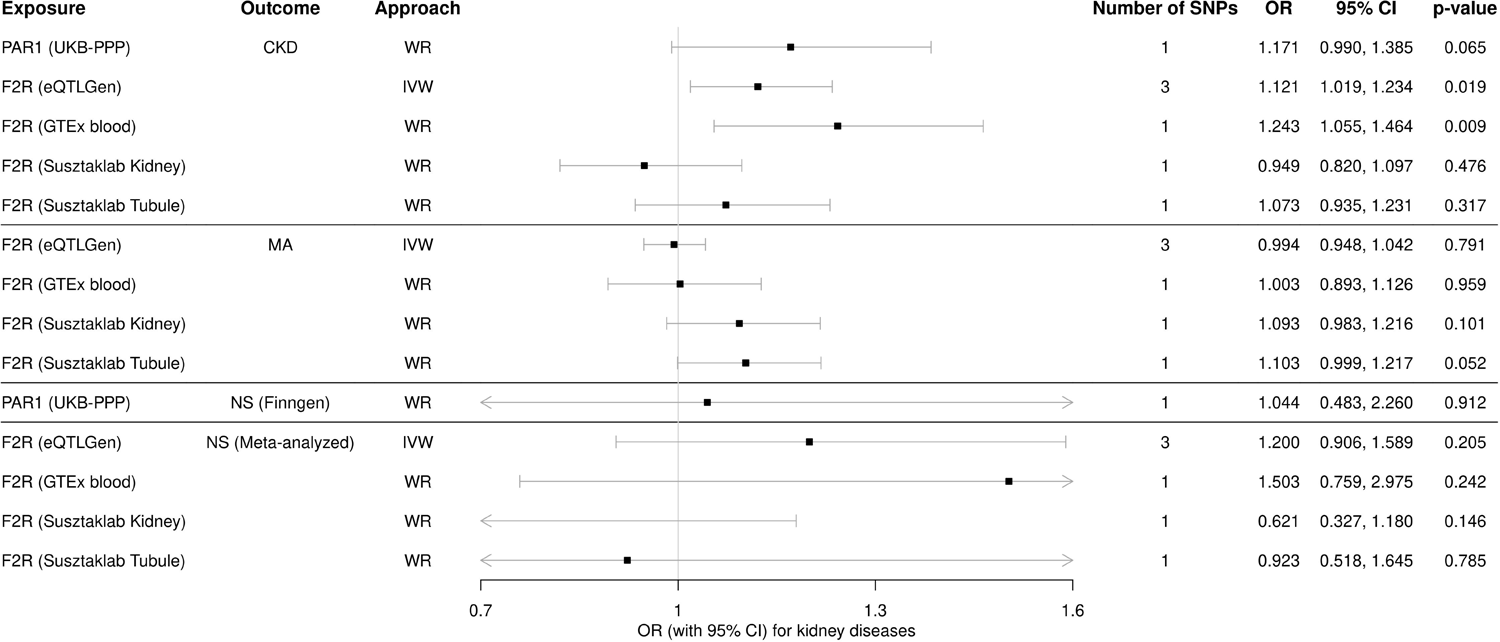
Results from MR analyses to estimate the causal effects of genetically-proxied lower plasma PAR1 level and *F2R* expression on kidney diseases. *F2R* is the gene name encoding the PAR1 protein. To avoid sample overlap issues, when using plasma PAR1 level from UKB-PPP, we used NS GWAS from FinnGen as the outcome, instead of meta-analyzed NS GWAS. UKB-PPP, UK Biobank Pharma Proteomics Project; eQTLGen, the eQTLGen Consortium; GTEx, The Genotype-Tissue Expression project; Susztaklab, Susztaklab Human Kidney eQTL Atlas; CKD, chronic kidney disease; MA, microalbuminuria; NS, nephrotic syndrome; WR, Wald ratio; IVW, inverse-variance-weighted; OR, odds ratios.

**Figure 5:**
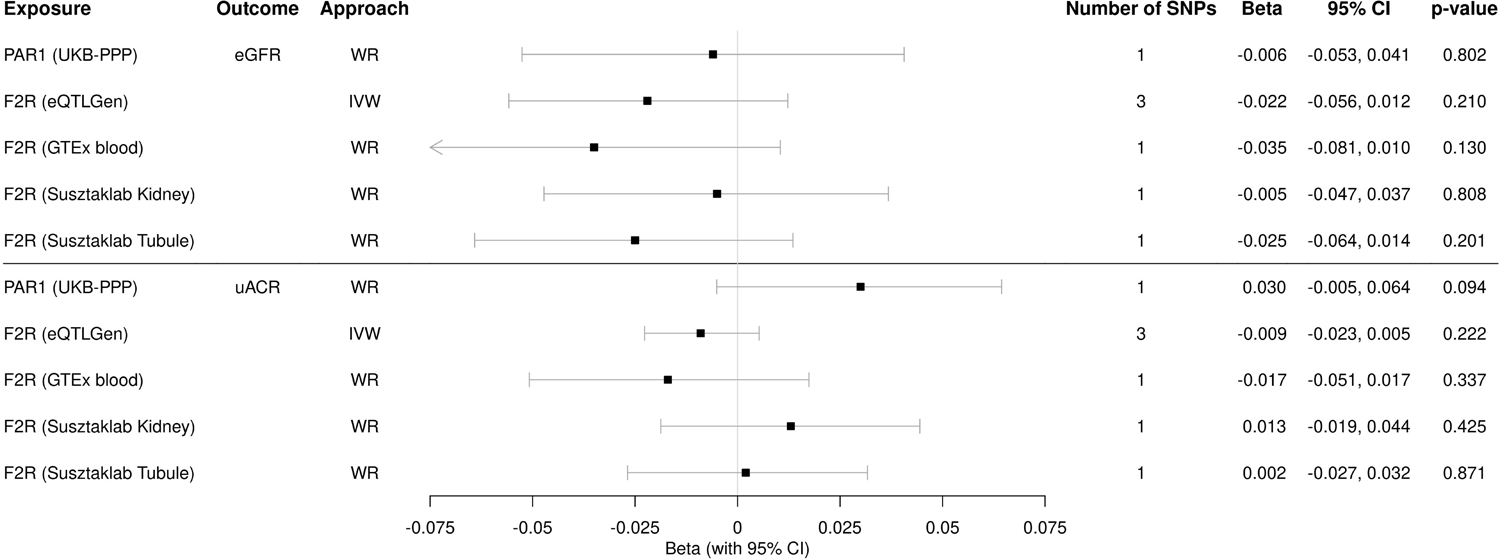
Results from MR analyses to estimate the causal effects of genetically-proxied lower PAR1 level on continuous renal phenotypes. The estimates are beta for log(eGFR) or log(uACR) per SD unit change in lower genetically-proxied PAR1 levels. UKB-PPP, UK Biobank Pharma Proteomics Project; eQTLGen, the eQTLGen Consortium; GTEx, The Genotype-Tissue Expression project; Susztaklab, Susztaklab Human Kidney eQTL Atlas; eGFR, estimated glomerular filtration rate; uACR, urinary albumin-to-creatinine ratio; WR, Wald ratio; IVW, inverse-variance-weighted.

The causal effects of genetically-proxied lower PAR1 or blood *F2R* on lower eGFR were *β* =-0.01, 95% CI: −0.05 to 0.04 in UKB-PPP; *β* =-0.02, 95% CI: −0.06 to 0.01 in eQTLGen; *β* =-0.04, 95% CI: −0.08 to 0.01 in GTEx. Similarly, the causal effects of genetically-proxied lower kidney *F2R* on eGFR were *β* =-0.01, 95% CI: −0.05 to 0.04 in Kidney; *β* =-0.03, 95% CI: −0.06 to 0.01 in Tubule. We also found no causal effects of PAR1, blood, or kidney *F2R* on uACR (e.g. eQTLGen: *β* =-0.01, 95% CI: −0.02 to 0.01).

### Sensitivity analyses results

#### Genetically-proxied lower *F2R* on serum albumin

We found inconsistent results of causal effects of genetically-proxied lower blood and kidney *F2R* on serum albumin (GTEx: *β* =-0.08, 95% CI: −0.12 to −0.04; kidney: *β* =0.05, 95% CI: 0.02 to 0.09) while other exposures showed inconclusive results (eQLTGen: *β* =-0.02, 95% CI: −0.05 to 0.01; Tubule: *β* =-0.02, 95% CI: −0.05 to 0.02) (Figure S2 and ST 4).

#### MR accounting for correlated instruments

We identified six and 13 weakly associated instruments from UKB-PPP and eQTLGen respectively, using LD clumping with an *r*^2^ threshold of 0.1. The other instruments did not change when using the less conservative *r*^2^ threshold (shown in ST 1).

For our positive control outcomes, the results of MR accounting for weakly correlated instruments were consistent with the main analyses (VTE, UKB-PPP: OR=0.80, 95% CI: 0.70 to 0.92 and eQTLGen: OR=0.95, 95% CI: 0.88 to 1.02; DVT, UKB-PPP: OR=0.67, 95% CI: 0.52 to 0.85 and eQTLGen: OR=0.85, 95% CI: 0.77 to 0.95) (Figure S3).

Genetically-proxied lower blood *F2R* increased CKD risk (eQTLGen: OR=1.08, 95% CI: 1.00 to 1.16) and decreased eGFR (*β* =-0.02, 95% CI: −0.04 to 0.00) (Figure S4, S5 and ST 5). Other results from the MR accounting for correlated instruments were inconclusive.

## Discussion

In this study, we found that genetically-proxied lower plasma PAR1 levels and *F2R* expression in blood tissue may increase CKD risk. This was not confirmed by analyses using *F2R* expression from kidney tissue. Evidence for the effect of genetically-proxied lower PAR1 on other renal phenotypes was also inconclusive. Our analyses of other renal phenotypes in kidney tissue also yield conflicting results, suggesting that genetically-proxied lower *F2R* in kidney tissue both increased serum albumin and elevated risk of MA while genetically-proxied lower blood *F2R* decreased albumin.

Vorapaxar is licensed as a treatment to reduce thrombotic cardiovascular events in individuals with prior myocardial infarction (MI) or peripheral arterial disease (PAD).^48^ In the RCTs (e.g. clinicaltrials.gov ID: NCT03207451 and NCT02545933), vorapaxar protected against the primary endpoint of cardiovascular mortality, MI, or stroke compared to the placebo (Hazard ratio = 0.87, p *<* 0.001).^1^ However, to our knowledge, there is currently no available GWAS of thrombotic cardiovascular events in individuals with prior MI or PAD. Vorapaxar works by inhibiting platelet aggregation induced by thrombin and thrombin receptor agonist peptide (TRAP). Therefore, we chose the platelet-aggregation-related phenotypes: VTE, DVT, and AET as positive control outcomes and found some evidence of causal effects of genetically-proxied lower PAR1 in blood and kidney tissues on lower risks of both VTE and DVT, as expected. The underpowered evidence for AET might be due to the small number of cases. There have been numerous experiments both *in vitro* and *in vivo* investigating the harmful effects of PAR1 activation on podocytes in NS and diabetic nephropathy^2, 49, 50^ and indicating some potential pathways mediated by TRPC6^2, 51^ and podocin.^51–53^ However, due to the limited number of NS cases, we found inconclusive evidence of the causal effects of plasma PAR1, blood *F2R* expression, and kidney *F2R* expression on NS.

There are several strengths of this study. First, we used multiple sources of evidence for genetic effects on PAR1, including molecular QTL from plasma, blood, and kidney (disease-related) tissues. We tested the colocalization of our instruments with eQTLs from the NS-disease-specific cohort and confirmed that our eQTL instruments from blood do colocalize with eQTL signals from the tubulointerstitial tissue of NS patients, under the assumption of a single causal variant in the *F2R* gene. Second, we employed thrombotic diseases as positive control outcomes to verify that the effect direction of the genetically-proxied lower PAR1 is consistent with the vorapaxar effect of PAR1 inhibition. Third, we used different kidney diseases and biomarkers as outcomes. We also applied an MR method that could account for correlated genetic instruments as a sensitivity analysis. Most of our findings were consistent, except those using the kidney tissue instruments, which showed opposing directions on albumin and MA. Finally, we conducted a novel NS GWAS in the UK Biobank European population, subsequently meta-analyzing it with the NS GWAS data from FinnGen Release 10. This represents the largest adulthood NS GWAS known to us to date. Despite this, the precision of our MR estimates for NS is too low to draw definitive conclusions.

This study also has limitations. First, most drugs typically do not influence target protein expression but rather bind to their target.^54^ For example, no clinical data shows that vorapaxar intake would change the PAR1 expression level in any cell type. However, in our drug-target MR, we instrumented genetically-proxied lower plasma-derived PAR1 protein level and genetically lower PAR1 blood- and kidney-derived mRNA expression. We assumed that genetically lower PAR1 expression would reduce the availability of receptors, and potentially proxy the effect of PAR1 inhibition. We also assumed that this proxy could represent PAR1 inhibition on the membrane of platelets (positive control) and podocytes (anti-PAR1 clinical trial). However, these assumptions may be invalid. pQTLs from UKB-PPP may not truly reflect PAR1 activity, because these data represent plasma-soluble PAR1 rather than functional transmembrane receptors.^55^ Second, most cell types express more than one protease-activated receptor (PAR) and the dimerization of PARs is common, for example, PAR1-PAR4 can form a dimer on the human platelet membrane. PAR1, PAR2 and PAR3 are encoded by the closely co-located genes *F2R*, *F2RL1*, and *F2RL2*. Data from the Open Targets platform (https://www.opentargets.org/) suggests that our PAR1 instruments may be responsible for more than one of the PAR-encoding genes. PAR1-PAR1, PAR1-PAR3, and PAR3-PAR3 dimerizations have all been found in transfected human embryonic kidney 293 (HEK293) cells, and PAR1-PAR3 heterodimer has also been found in murine podocytes. These dimerizations, plus the genomic proximity of the genes, may have introduced pleiotropy into our analyses, which will have affected the MR results for both the positive control and kidney-related outcomes. Third, we could not apply traditional MR sensitivity analyses, such as MR-Egger,^45^ weighted median,^56^ and weighted mode^57^ methods, due to the small number of genetic instruments. Fourth, our MR results represent the life-long effects of lower PAR-1, which is not equivalent to taking an anti-PAR-1 treatment, which would be over a much shorter period. Therefore, the effect estimates of our MR findings may not match drug effects documented in clinical trials. Finally, our MR analyses used data from European ancestry individuals, leading to large sample sizes at the expense of generalisability. Despite this, the number of NS cases is still limited, leading to the low precision of these results.

In conclusion, our study shows some evidence that genetically-proxied lower PAR1 in plasma and *F2R* expression in blood and kidney tissues reduces the risks of VTE and DVT. Genetically-proxied lower *F2R* expression levels in blood tissue may increase the risk of CKD, but effects on NS and MA were inconclusive. We also found inconsistent effects of blood and kidney *F2R* on serum albumin. Regular monitoring of renal function (e.g. eGFR, uACR, and albumin) in patients on vorapaxar is advisable to detect early signs of CKD and understand the potential heterogeneity of PAR1 tissue-specific effects. More extensive studies are needed to determine the effects of PAR1 inhibition on NS and to explore the mechanisms underlying the observed increase in CKD risk.

## Supporting information

Not applicable

## Funds

This work was supported by the UK Medical Research Council Integrative Epidemiology Unit (MC_UU_00032/03).

## Author Contributions

HT conducted the analyses and drafted the manuscript. VMW proposed the potential of MR studies on PAR-1. VMW and TRG equally contributed to the supervision and manuscript review. All authors approved the final version for submission.

## Data available

The summary-level data from the Neale lab (http://www.nealelab.is/uk-biobank/) at OpenG-WAS (https://gwas.mrcieu.ac.uk/), CKDGen (http://ckdgen.imbi.uni-freiburg.de/), UKB-PPP (https://metabolomips.org/ukbbpgwas/), Susztaklab Human Kidney eQTL,(https://susztaklab.com/Kidney_eQTL/), GTEx (https://www.gtexportal.org/home/) and NEP-TUNE are respectively publicly available. The Genotype-Tissue Expression (GTEx) Project was supported by the Common Fund of the Office of the Director of the National Institutes of Health, and by NCI, NHGRI, NHLBI, NIDA, NIMH, and NINDS. The data used for the analyses described in this manuscript were obtained from the GTEx Portal on 16/04/2024.

## References

1. Morrow DA, Braunwald E, Bonaca MP, et al. Vorapaxar in the Secondary Prevention of Atherothrombotic Events. New England Journal of Medicine. 2012;366(15):1404–1413. DOI: 10.1056/nejmoa1200933.

2. May CJ, Chesor M, Hunter SE, et al. Podocyte protease activated receptor 1 stimulation in mice produces focal segmental glomerulosclerosis mirroring human disease signaling events. Kidney International. 2023;104(2):265–278. DOI: 10.1016/j.kint.2023.02.031.

3. Ginsberg JM, Chang BS, Matarese RA, Garella S. Use of Single Voided Urine Samples to Estimate Quantitative Proteinuria. New England Journal of Medicine. 1983;309(25):1543–1546. DOI: 10.1056/nejm198312223092503.

4. Orth SR, Ritz E. The Nephrotic Syndrome. New England Journal of Medicine. 1998;338(17):1202–1211. DOI: 10.1056/nejm199804233381707.

5. Rosenberg AZ, Kopp JB. Focal Segmental Glomerulosclerosis. Clinical Journal of the American Society of Nephrology. 2017;12(3):502–517. DOI: 10.2215/cjn.05960616.

6. Webster AC, Nagler EV, Morton RL, Masson P. Chronic Kidney Disease. The Lancet. 2017;389(10075):1238–1252. DOI: 10.1016/s0140-6736(16)32064-5.

7. Kalantar-Zadeh K, Jafar TH, Nitsch D, Neuen BL, Perkovic V. Chronic kidney disease. The Lancet. 2021;398(10302):786–802. DOI: 10.1016/s0140-6736(21)00519-5.

8. Stevens PE, Ahmed SB, Carrero JJ, et al. KDIGO 2024 Clinical Practice Guideline for the Evaluation and Management of Chronic Kidney Disease. Kidney International. 2024;105(4):S117–S314. DOI: 10.1016/j.kint.2023.10.018.

9. Lamb EJ, McTaggart MP, Stevens PE. Why albumin to creatinine ratio should replace protein to creatinine ratio: it is not just about nephrologists. Annals of Clinical Biochemistry: International Journal of Laboratory Medicine. 2013;50(4):301–305. DOI: 1.1177/0004563212473284.

10. Kalluri R. Proteinuria with and without Renal Glomerular Podocyte Effacement. Journal of the American Society of Nephrology. 2006;17(9):2383–2389. DOI: 10.1681/asn.2006060628.

11. Chang DR, Yeh HC, Ting IW, et al. The ratio and difference of urine protein-to-creatinine ratio and albumin-to-creatinine ratio facilitate risk prediction of all-cause mortality. Scientific Reports. 2021;11(1). DOI: 10.1038/s41598-021-86541-3.

12. Davey-Smith G. Data dredging, bias, or confounding. BMJ. 2002;325(7378):1437–1438. DOI: 10.1136/bmj.325.7378.1437.

13. Lawlor DA, Harbord RM, Sterne JAC, Timpson N, Davey-Smith G. Mendelian randomization: Using genes as instruments for making causal inferences in epidemiology. Statistics in Medicine. 2008;27(8):1133–1163. DOI: 10.1002/sim.3034.

14. Davey-Smith G, Hemani G. Mendelian randomization: genetic anchors for causal inference in epidemiological studies. Human Molecular Genetics. 2014;23(R1):R89–R98. DOI: 10.1093/hmg/ddu328.

15. Swerdlow DI, Preiss D, Kuchenbaecker KB, et al. HMG-coenzyme A reductase inhibition, type 2 diabetes, and bodyweight: evidence from genetic analysis and randomised trials. The Lancet. 2015;385(9965):351–361. DOI: 10.1016/s0140-6736(14)61183-1.

16. Schmidt AF, Swerdlow DI, Holmes MV, et al. PCSK9 genetic variants and risk of type 2 diabetes: a mendelian randomisation study. The Lancet Diabetes amp; Endocrinology. 2017;5(2):97–105. DOI: 10.1016/s2213-8587(16)30396-5.

17. Walker VM, Davey-Smith G, Davies NM, Martin RM. Mendelian randomization: a novel approach for the prediction of adverse drug events and drug repurposing opportunities. International Journal of Epidemiology. 2017;46(6):2078–2089. DOI: 10.1093/ije/dyx207.

18. Gill D, Georgakis MK, Walker VM, et al. Mendelian randomization for studying the effects of perturbing drug targets. Wellcome Open Research. 2021;6:16. DOI: 10.12688/wellcomeopenres.16544.2.

19. Fang S, Yarmolinsky J, Gill D, et al. Association between genetically proxied PCSK9 inhibition and prostate cancer risk: A Mendelian randomisation study. PLOS Medicine. 2023;20(1):e1003988. DOI: 10.1371/journal.pmed.1003988.

20. Ference BA, Majeed F, Penumetcha R, Flack JM, Brook RD. Effect of Naturally Random Allocation to Lower Low-Density Lipoprotein Cholesterol on the Risk of Coronary Heart Disease Mediated by Polymorphisms in NPC1L1, HMGCR, or Both. Journal of the American College of Cardiology. 2015;65(15):1552–1561. DOI: 10.1016/j.jacc.2015.02.020.

21. Ference BA, Robinson JG, Brook RD, et al. Variation inPCSK9andHMGCRand Risk of Cardiovascular Disease and Diabetes. New England Journal of Medicine. 2016;375(22):2144–2153. DOI: 10.1056/nejmoa1604304.

22. Zheng J, Haberland V, Baird D, et al. Phenome-wide Mendelian randomization mapping the influence of the plasma proteome on complex diseases. Nature Genetics. 2020;52(10):1122–1131. DOI: 10.1038/s41588-020-0682-6.

23. Skrivankova VW, Richmond RC, Woolf BAR, et al. Strengthening the Reporting of Observational Studies in Epidemiology Using Mendelian Randomization: The STROBE-MR Statement. JAMA. 2021;326(16):1614. DOI: 10.1001/jama.2021.18236.

24. Sun BB, Chiou J, Traylor M, et al. Plasma proteomic associations with genetics and health in the UK Biobank. Nature. 2023;622(7982):329–338. DOI: 10.1038/s41586-023-06592-6.

25. Võsa U, Claringbould A, Westra HJ, et al. Large-scale cis- and trans-eQTL analyses identify thousands of genetic loci and polygenic scores that regulate blood gene expression. Nature Genetics. 2021;53(9):1300–1310. DOI: 10.1038/s41588-021-00913-z.

26. Aguet F, Anand S, Ardlie KG, et al. The GTEx Consortium atlas of genetic regulatory effects across human tissues. Science. 2020;369(6509):1318–1330. DOI: 10.1126/science.aaz1776.

27. Liu H, Doke T, Guo D, et al. Epigenomic and transcriptomic analyses define core cell types, genes and targetable mechanisms for kidney disease. Nature Genetics. 2022;54(7):950–962. DOI: 10.1038/s41588-022-01097-w.

28. Sheng X, Guan Y, Ma Z, et al. Mapping the genetic architecture of human traits to cell types in the kidney identifies mechanisms of disease and potential treatments. Nature Genetics. 2021;53(9):1322–1333. DOI: 10.1038/s41588-021-00909-9.

29. Han SK, McNulty MT, Benway CJ, et al. Mapping genomic regulation of kidney disease and traits through high-resolution and interpretable eQTLs. Nature Communications. 2023;14(1). DOI: 10.1038/s41467-023-37691-7.

30. Kurki MI, Karjalainen J, Palta P, et al. FinnGen provides genetic insights from a well-phenotyped isolated population. Nature. 2023;613(7944):508–518. DOI: 10.1038/s41586-022-05473-8.

31. Wuttke M, Li Y, Li M, et al. A catalog of genetic loci associated with kidney function from analyses of a million individuals. Nature Genetics. 2019;51(6):957–972. DOI: 10.1038/s41588-019-0407-x.

32. Teumer A, Li Y, Ghasemi S, et al. Genome-wide association meta-analyses and fine-mapping elucidate pathways influencing albuminuria. Nature Communications. 2019;10(1). DOI: 10.1038/s41467-019-11576-0.

33. Willer CJ, Li Y, Abecasis GR. METAL: fast and efficient meta-analysis of genomewide association scans. Bioinformatics. 2010;26(17):2190–2191. DOI: 10.1093/bioinformatics/btq340.

34. Elsworth B, Lyon M, Alexander T, et al. The MRC IEU OpenGWAS data infrastructure. 2020. DOI: 10.1101/2020.08.10.244293.

35. Gadegbeku CA, Gipson DS, Holzman LB, et al. Design of the Nephrotic Syndrome Study Network (NEPTUNE) to evaluate primary glomerular nephropathy by a multidisciplinary approach. Kidney international. 2013;83(4):749–756.

36. Hemani G, Elsworth B, Palmer T, Rasteiro R. *ieugwasr: Interface to the ‘OpenGWAS’ Database API* 2024. R package version 0.2.2–9000, https://mrcieu.github.io/ieugwasr/.

37. Burgess S, Thompson SG. Avoiding bias from weak instruments in Mendelian randomization studies. International Journal of Epidemiology. 2011;40(3):755–764. DOI: 10.1093/ije/dyr036.

38. Davies NM, Holmes MV, Davey-Smith G. Reading Mendelian randomisation studies: a guide, glossary, and checklist for clinicians. BMJ. 2018:k601. DOI: 10.1136/bmj.k601.

39. Hemani G, Zheng J, Elsworth B, et al. The MR-Base platform supports systematic causal inference across the human phenome. eLife. 2018;7. DOI: 10.7554/elife.34408.

40. Giambartolomei C, Vukcevic D, Schadt EE, et al. Bayesian Test for Colocalisation between Pairs of Genetic Association Studies Using Summary Statistics. PLoS Genetics. 2014;10(5):e1004383. DOI: 10.1371/journal.pgen.1004383.

41. Burgess S, Davies NM, Thompson SG. Bias due to participant overlap in two-sample Mendelian randomization. Genetic Epidemiology. 2016;40(7):597–608. DOI: 10.1002/gepi.21998.

42. Wald A. The Fitting of Straight Lines if Both Variables are Subject to Error. The Annals of Mathematical Statistics. 1940;11(3):284–300. DOI: 10.1214/aoms/1177731868.

43. Burgess S, Small DS, Thompson SG. A review of instrumental variable estimators for Mendelian randomization. Statistical Methods in Medical Research. 2015;26(5):2333–2355. DOI: 10.1177/0962280215597579.

44. Burgess S, Butterworth A, Thompson SG. Mendelian Randomization Analysis With Multiple Genetic Variants Using Summarized Data. Genetic Epidemiology. 2013;37(7):658–665. DOI: 10.1002/gepi.21758.

45. Bowden J, Davey-Smith G, Burgess S. Mendelian randomization with invalid instruments: effect estimation and bias detection through Egger regression. International Journal of Epidemiology. 2015;44(2):512–525. DOI: 10.1093/ije/dyv080.

46. Lang J, Katz R, Ix JH, et al. Association of serum albumin levels with kidney function decline and incident chronic kidney disease in elders. Nephrology Dialysis Transplantation. 2017;33(6):986–992. DOI: 10.1093/ndt/gfx229.

47. Burgess S, Zuber V, Valdes-Marquez E, Sun BB, Hopewell JC. Mendelian randomization with fine-mapped genetic data: Choosing from large numbers of correlated instrumental variables. Genetic Epidemiology. 2017;41(8):714–725. DOI: 10.1002/gepi.22077.

48. Poole RM, Elkinson S. Vorapaxar: First Global Approval. Drugs. 2014;74(10):1153–1163. DOI: 10.1007/s40265-014-0252-2.

49. May CJ, Welsh GI, Chesor M, et al. Human Th17 cells produce a soluble mediator that increases podocyte motility via signaling pathways that mimic PAR-1 activation. American Journal of Physiology-Renal Physiology. 2019;317(4):F913–F921. DOI: 10.1152/ajprenal.00093.2019.

50. Bohovyk R, Khedr S, Levchenko V, et al. Protease-Activated Receptor 1–Mediated Damage of Podocytes in Diabetic Nephropathy. Diabetes. 2023;72(12):1795–1808. DOI: 10.2337/db23-0032.

51. Kim EY, Roshanravan H, Dryer SE. Changes in podocyte TRPC channels evoked by plasma and sera from patients with recurrent FSGS and by putative glomerular permeability factors. Biochimica et Biophysica Acta (BBA) - Molecular Basis of Disease. 2017;1863(9):2342–2354. DOI: 10.1016/j.bbadis.2017.06.010.

52. Harris JJ, McCarthy HJ, Ni L, et al. Active proteases in nephrotic plasma lead to a podocin-dependent phosphorylation of <scp>VASP</scp> in podocytes via protease activated receptor-1. The Journal of Pathology. 2013;229(5):660–671. DOI: 10.1002/path.4149.

53. Anderson M, Kim EY, Hagmann H, Benzing T, Dryer SE. Opposing effects of podocin on the gating of podocyte TRPC6 channels evoked by membrane stretch or diacylglycerol. American Journal of Physiology-Cell Physiology. 2013;305(3):C276–C289. DOI: 10.1152/ajpcell.00095.2013.

54. Isik Z, Baldow C, Cannistraci CV, Schroeder M. Drug target prioritization by perturbed gene expression and network information. Scientific Reports. 2015;5(1). DOI: 10.1038/srep17417.

55. Kremers BMM, Posma JN, Heitmeier S, et al. Discovery of four plasmatic biomarkers potentially predicting cardiovascular outcome in peripheral artery disease. Scientific Reports. 2022;12(1). DOI: 10.1038/s41598-022-23260-3.

56. Bowden J, Davey-Smith G, Haycock PC, Burgess S. Consistent Estimation in Mendelian Randomization with Some Invalid Instruments Using a Weighted Median Estimator. Genetic Epidemiology. 2016;40(4):304–314. DOI: 10.1002/gepi.21965.

57. Hartwig FP, Davey-Smith G, Bowden J. Robust inference in summary data Mendelian randomization via the zero modal pleiotropy assumption. International Journal of Epidemiology. 2017;46(6):1985–1998. DOI: 10.1093/ije/dyx102.

