## Supplementary Materials for "Genetically-proxied lower PAR1 and renal phenotypes: a drug-target Mendelian Randomization study"

Corresponding Author:

Mr. Haotian Tang

Medical Research Council Integrative Epidemiology Unit, Bristol Medical School, University of Bristol, Oakfield House, Oakfield Grove, Bristol, BS8 2BN, United Kingdom.

[*]MRC Integrative Epidemiology Unit, Bristol Medical School, University of Bristol, Bristol, UK

[†]VMW and TRG equally contributed to the supervision and manuscript review.

### GWAS in UKB

UKB participants with the ICD-10 code N04 were considered as NS cases and UKB participants with none of the codes N00 to N08 were considered as controls (followed by definition from FinnGen Risteys R10, available from `https://r10.risteys.finregistry.fi/endpoints/N14_NEPHROTICSYND`). We performed NS GWAS in the UKB European population (349 cases and 459,687 controls, by using the UKB GWAS pipeline which was developed by the Medical Research Council (MRC) Integrative Epidemiology Unit (IEU).[1] We applied the linear mixed modeling approach, BOLT-LMM, to account for population stratification and relatedness. The covariates in the model included genotyping chip, sex, and age.

### Software and code availability

Analyses were conducted using R v4.2.2. All LD clumping for genetic instruments was conducted using the `ieugwasr` v1.0.0[2] R package (`https://mrcieu.github.io/ieugwasr/`). Colocalization analyses were performed using `coloc`[3] v5.2.3 R package (`https://chr1swallace.github.io/coloc/articles/a01_intro.html`) and the colocalization evidence were visualized by using the `locuszoomr` R v0.3.0 package (`https://github.com/myles-lewis/locuszoomr`). MR analyses with independent instruments were performed with the `TwoSampleMR`[4] v0.6.1 R package (`https://github.com/MRCIEU/TwoSampleMR`) and MR analyses accounting for correlated instruments were conducted with the `MendelianRandomization` v0.10.0 R package (available in the Comprehensive R Archive Network). Chromosome and base-pair position of NephQTL2 GWAS were matched with RSID by using `MungeSumstats`[5] v1.10.1 R package (`https://github.com/neurogenomics/MungeSumstats`). Forest plots were generated using the `forestplot` v3.1.3 R package (`https://github.com/gforge/forestplot`). All scripts are available from GitHub (`https://github.com/Haotian2020/PAR1_MRdrugtarget_Project`).

### Supplementary Figures

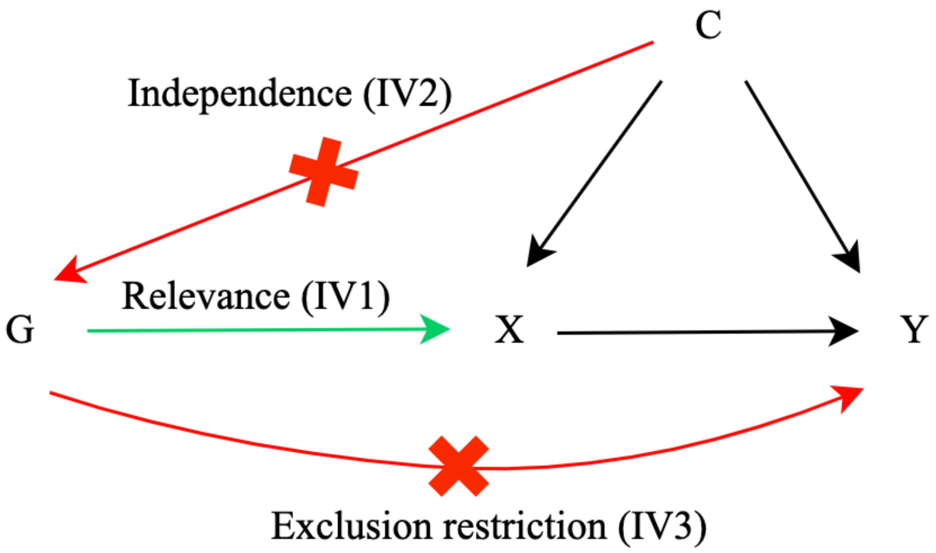


Figure S1: Directed acyclic graph representing the three core assumptions for Mendelian Randomization analyses: relevance independence and exclusion restriction. G, genetic instruments; X, exposure; Y, outcome; C, confounder.

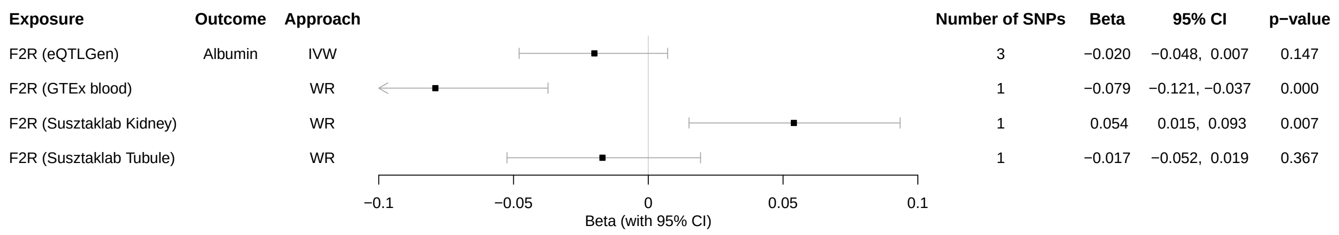


Figure S2: Results from MR analyses to estimate the associations between PAR1 level and other kidney function biomarkers and diseases. *F2R* is the gene encoding the PAR1 protein. UKB-PPP, UK Biobank Pharma Proteomics Project; eQTLGen, the eQTLGen Consortium; GTEx, The Genotype-Tissue Expression project; Susztaklab, Susztaklab Human Kidney eQTL Atlas; WR, Wald ratio; IVW, inverse-variance-weighted.

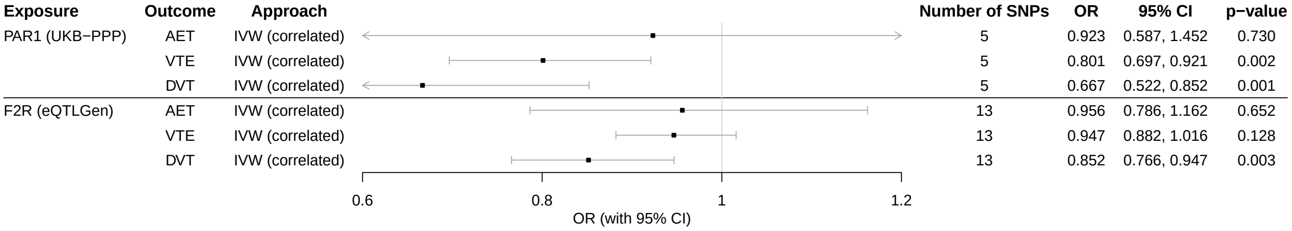


Figure S3: Results from MR analyses accounting for correlated instruments to estimate the causal effects of plasma PAR1 and blood *F2R* expression levels on thrombotic diseases. UKB-PPP, UK Biobank Pharma Proteomics Project; eQTLGen, the eQTLGen Consortium; ATE, Arterial thromboembolism; VTE, venous thromboembolism; DVT, deep vein thrombosis; IVW, inverse-variance-weighted.

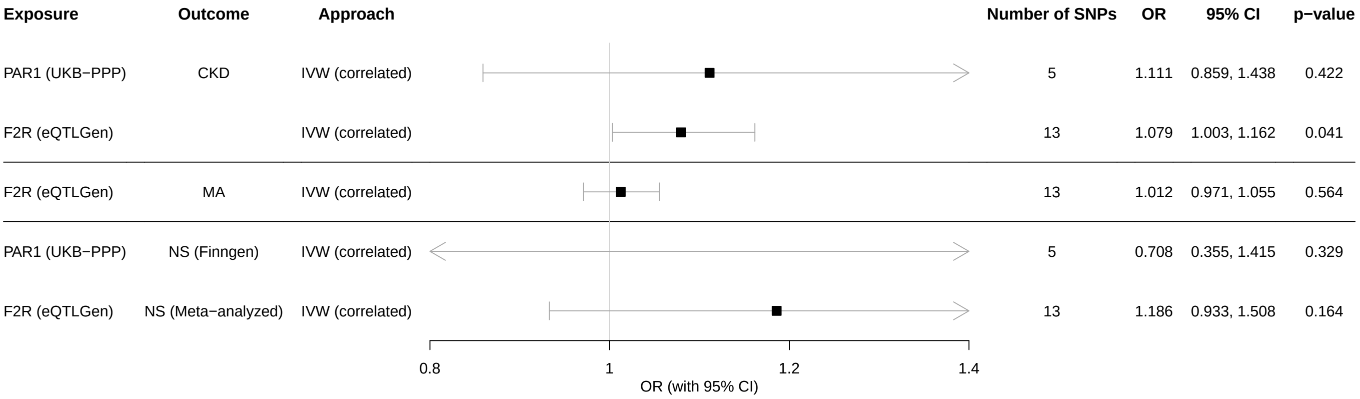


Figure S4: Results from MR analyses accounting for correlated instruments to estimate the causal effects of genetically-proxied plasma PAR1 and blood *F2R* expression level on kidney diseases. *F2R* is the gene name encoding the PAR1 protein. To avoid sample overlap issues, when using plasma PAR1 level from UKB-PPP, we used NS GWAS from FinnGen as the outcome, instead of meta-analyzed NS GWAS. UKB-PPP, UK Biobank Pharma Proteomics Project; eQTLGen, the eQTLGen Consortium; GTEx, The Genotype-Tissue Expression project; Susztaklab, Susztaklab Human Kidney eQTL Atlas; CKD, chronic kidney disease; MA, microalbuminuria; NS, nephrotic syndrome; IVW, inverse-variance-weighted.

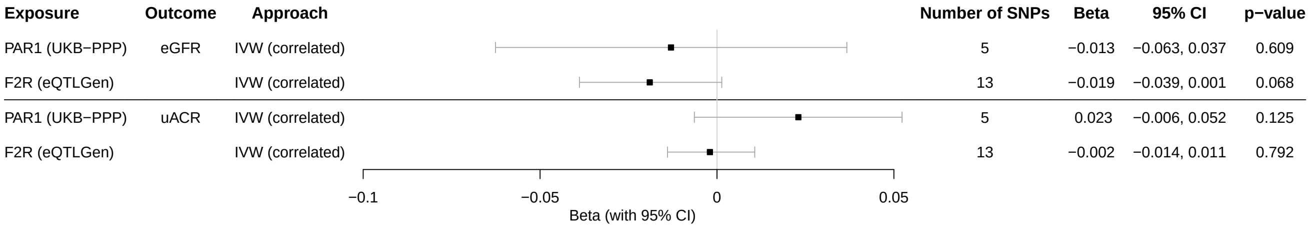


Figure S5: Results from MR analyses accounting for correlated instruments to estimate the causal effects of plasma PAR1 level and blood *F2R* expression on kidney function biomarkers. *F2R* is the gene name encoding the PAR1 protein. UKB-PPP, UK Biobank Pharma Proteomics Project; eQTLGen, the eQTLGen Consortium; eGFR, estimated glomerular filtration rate; uACR, urinary albumin-creatinine ratio; IVW, inverse-variance-weighted.

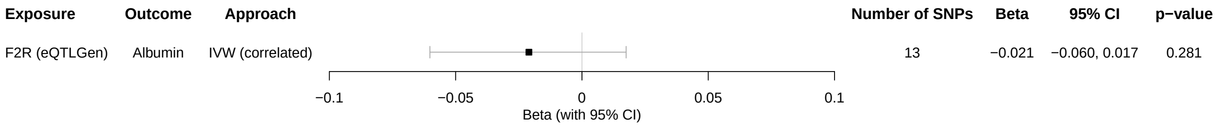


Figure S6: Results from MR analyses accounting for correlated instruments to estimate the causal effects of genetically-proxied blood *F2R* expression level on serum albumin. *F2R* is the gene name encoding the PAR1 protein. eQTLGen, the eQTLGen Consortium; IVW, inverse-variance-weighted.

### References

1. Mitchell R, Hemani G, Dudding T, Corbin L, Harrison S, Paternoster L. UK Biobank Genetic Data: MRC-IEU Quality Control, version 2. *University of Bristol.* 2019.

2. Hemani G, Elsworth B, Palmer T, Rasteiro R. *ieugwasr: Interface to the 'OpenGWAS' Database API* 2024. R package version 0.2.2-9000, https://mrcieu.github.io/ieugwasr/.

3. Giambartolomei C, Vukcevic D, Schadt EE, *et al.* Bayesian Test for Colocalisation between Pairs of Genetic Association Studies Using Summary Statistics. *PLoS Genetics.* 2014;10(5):e1004383. DOI: 10.1371/journal.pgen.1004383.

4. Hemani G, Zheng J, Elsworth B, *et al.* The MR-Base platform supports systematic causal inference across the human phenome. *eLife.* 2018;7. DOI: 10.7554/elife.34408.

5. Murphy AE, Schilder BM, Skene NG. MungeSumstats: a Bioconductor package for the standardization and quality control of many GWAS summary statistics. *Bioinformatics.* 2021;37(23):4593–4596. DOI: 10.1093/bioinformatics/btab665.
